# The phenomenology of tics and tic-like behavior in TikTok

**DOI:** 10.1101/2021.09.08.21263218

**Authors:** Alonso Zea Vera, Adrienne Bruce, Jordan Garris, Laura Tochen, Poonam Bhatia, Rebecca K Lehman, Wendi Lopez, Steve W. Wu, Donald L. Gilbert

**Affiliations:** Division of Neurology, Cincinnati Children’s Hospital Medical Center, Cincinnati, OH, USA; Department of Neurology, Children’s National Hospital, Washington, DC, USA; Department of Pediatrics, Prisma Health, Greenville, SC, USA; University of South Carolina School of Medicine Greenville, Greenville, SC, USA; Department of Neurology, University of Virginia Medical Center, Charlottesville, VA, USA; Pediatric Movement Disorders Program, Barrow Neurological Institute, Phoenix Children’s Hospital, Phoenix, AZ, USA; Prisma Health-Midlands, Columbia, SC, USA; Department of Neurology, University of South Carolina, Columbia, SC, USA; Division of Behavioral Medicine and Clinical Psychology, Cincinnati Children’s Hospital Medical Center, Cincinnati, OH, USA; Department of Pediatrics, University of Cincinnati College of Medicine, Cincinnati, OH, USA

**Keywords:** Tourette Syndrome, Tics, Functional tic-like disorder, Social Media

## Abstract

**Background and Objective:** Pediatricians and pediatric subspecialists worldwide have reported a marked increase in functional (conversion) disorders with tic-like behaviors during the COVID-19 pandemic. These patients often report frequent viewing of Tourette Syndrome (TS) TikTok videos, suggesting disease modeling. We aimed to evaluate tic phenomenology in videos posted on TikTok.

**Methods:** The 100 most-viewed videos under #tourettes in TikTok were randomly assigned to two primary reviewers (n=3; <2 years independent practice), all pediatric neurologists specializing in movement disorders, for extraction and classification of tic phenomenology. Initial disagreements were solved by consensus. If not resolved, a senior reviewer (n=5; >2 years independent pediatric movement-disorder practice) served as tiebreaker. In addition, two primary and one senior reviewer rated each video on a Likert scale from 1= “All the tics are typical of TS” to 5= “None of the tics are typical of TS”. Mean scores and Spearman correlation between primary and senior reviewers were calculated.

**Results:** Six videos without tic-like behaviors were excluded. Most videos depicted coprophenomena (coprolalia: 53.2%; copropraxia: 20.2%), often with unusual characteristics. Frequently, videos demonstrated atypical phenomenology such as very strong influence by the environment (motor: 54.3%; phonic: 54.3%), aggression (19.1%), throwing objects (22.3%), self-injurious behaviors (27.7%), and long phrases (>3 words; 45.7%). Most videos portrayed atypical, non-tic behaviors (Median [IQR] Likert ratings: Primary 4.5 [4-5]; Senior 5 [3-5]). Primary vs. senior rater scores demonstrated moderate agreement (r = 0.46; p<0.001).

**Conclusions:** TS symptoms portrayals on highly-viewed TikTok videos are predominantly not representative or typical of TS.

**Highlights:**

- Many teenagers with functional tic-like disorder have been reported during the COVID-19 pandemic.
- These patients report increased viewing of Tourette Syndrome TikTok videos, a popular social media platform, and present with similar tic-like behaviors.
- Current TikTok videos are poorly representative of Tourette syndrome and more consistent with functional tic-like behavior.
- We provide a detailed description of the phenomenology tics and tic-like behavior portrayed in TikTok.

## INTRODUCTION

Tourette Syndrome (TS) affects nearly 1% of the population and is characterized by persistent motor and phonic tics.^1^ TS most commonly presents in boys and has a gradual onset, with tics appearing predominantly before the age of 10 years. Tics are brief, repetitive, patterned, non-rhythmic movements or vocalizations, sometimes performed in response to premonitory urges. Tics are divided in simple (e.g., blinking, grunting) and complex (e.g., gestures, words and phrases) Most tics have no clear external triggers and appear out of context.^1^ Generally, the emergence of tics follows a rostrocaudal distribution. Concentration, focusing on an activity or performing purposeful actions in the same muscles commonly decreases tics, limiting their interference with voluntary actions. Most TS patients can voluntarily suppress their tics, albeit only momentarily.^1^

Functional tic-like disorder (FTLD) is a subtype of functional movement disorder characterized by tic-like behaviors – sounds or movements that resemble tics. In contrast to TS, FTLD more commonly presents with multiple symptoms in a short time interval in adolescent and young adult females.^2^ Compared to tics, functional tic-like behaviors involve the arms or trunk more than the face, interfere more with voluntary actions, are less suppressible, and are more suggestible.^2, 3^ FTLD can be seen in combination with other functional neurologic symptoms and has a poor response to common tic medications.^2^ FTLD can also emerge in persons with TS, and in some cases, phenomenological features overlap between TS and FTLD.^2^

A rise in tic-referrals to movement disorders clinics has occurred during the COVID-19 pandemic. TS patients report worse symptoms during the pandemic,^4^ possibly related to increased anxiety and confinement.^5^ In parallel, several groups have noticed a dramatic increase in teenagers, mainly females, presenting with sudden onset FTLD.^6-9^ Although a general increase in functional movement disorders has also been reported,^10^ there has been a disproportionate increase in FTLD. Until recently FTLD represented only a minority of functional movement disorders.^11^ This brings the question “Why, specifically, tics?”.^12^

The platform TikTok (www.tiktok.com) has become very popular among teenagers.^13^ The use of social media and the viewing of the #tourettes TikTok webpage has significantly increased during the pandemic.^6, 14^ The increase use of social media may have had a detrimental effect on mental health during the pandemic.^14^ Many patients with FTLD report watching TS videos or posting videos of their movements in TikTok. This suggests a potential association between TikTok and the rise in FTLD.^6-9^ The accuracy of medical videos in social media is highly variable. In a previous study, 66% of videos portraying movement disorders were qualified as functional by experts in the field.^15^ However, there is limited information on the characteristics of the current videos available in TikTok. The goal of this study is to characterize and evaluate the phenomenology of tics and tic-like behavior portrayed in TikTok.

## METHODS

### Search Strategy

Our video search was performed on a single day on March 27^th^, 2021. Before starting the search, the computer’s cache/cookies and search history were cleared to prevent the effects of previous searches on our sample. We looked for all the videos under #tourettes. We chose this hashtag because it is commonly used in videos portraying TS and recently had an increase in popularity.

We selected the 100 most-viewed videos. This number represents a reasonable sample of the videos that would be seen by a person searching for TS in TikTok while maintaining the feasibility of evaluating a large number of videos. Duplicated videos or videos that did not show tics were excluded. The number of comments and “likes” were recorded for each video. Of note, some of the videos portrayed the same subject at different times. We did not limit our search to only one video per subject because we wanted our sample to realistically represent all available videos in TikTok. We did not group the videos by the portrayed subject.

### Video review

All reviewers were pediatric neurologists specializing in movement disorders. Primary reviewers (AZV, AB, and JG) had less than 2 years of independent practice. Senior reviewers (SW, LT, RL, PB, and DG) were more experienced specialists. Based on a literature review and clinical experience, an initial list of common, atypical, and severe tic-phenomenology was generated for use in quantifying phenomenology in videos. We rated the presence or absence of each feature in the entire video, allowing for the possibility of multiple tics per video. A pilot was conducted with 10 videos. After analysis, primary reviewers discussed difficulties in data extraction. As a result, some tic-like characteristics judged too difficult to reliably and accurately assess in short and edited videos were eliminated. The final list of tic phenomenology and definitions used for data extraction is presented in the Supplementary Materials.

We utilized the construct “context-dependent tics” (CDT) in our data extraction to classify behaviors that appeared to be strongly influenced by the environment. This included two features that frequently occurred together and were difficult to assess separately based only on short videos. The first are behaviors that are very specific to the external situation of the subject. For example, involuntary phrases that followed the content of an ongoing conversation or complex motor behaviors using objects in the environment. The second are tics that are triggered by an external stimulus, such as clapping specifically when seeing a dog; these have been called stimulus-bound or reflex tics.^16^ Similar to other studies, we excluded ecophenomena (repeating other people’s words or actions) and paliphenomena (repeating one’s own words or actions) from this category.^16^

Each video was randomly assigned to two primary reviewers who independently quantified and extracted the characteristics of tics or tic-like behaviors, blinded to each other’s initial assessment. Both primary reviewers discussed disagreements and tried to come to a consensus. If this was not achieved, a senior reviewer was the tiebreaker. Any movement or sound that was portrayed as a tic and is likely to be interpreted as a tic by a lay audience was included. Qualitative descriptions of other patterns of tics in these videos were also obtained.

In addition to evaluating tic phenomenology, all videos were rated globally using a Likert scale from 1= “All of the movements or sounds portrayed are typical of a primary tic disorder such as Tourette syndrome” to 5= “None of the movements or sounds portrayed are typical of a primary tic disorder such as Tourette syndrome”. Each video was randomly assigned to be rated by 2 primary reviewers, who also evaluated tic phenomenology, and 1 senior reviewer, all of them blind to the others’ assessment.

### Ethical Considerations

All videos included were publicly accessible in social media, therefore considered part of the public domain. Consistent with other studies of publicly available material in social media, consent from creators and ethics approval were not obtained.^17, 18^ No identifiable information is included in the manuscript.

### Data Analysis

Tic phenomenology was summarized by percentages of videos representing each feature. In addition, qualitative descriptions are provided. Likert scale ratings of both primary reviewers were averaged for comparison with the senior reviewer. Likert scale ratings were summarized using medians and interquartile ranges. Correlation between mean primary reviewers’ score and senior reviewer’s score was calculated using Spearman correlation.

## RESULTS

### Videos

The 100 videos in the initial search were posted by 38 different users. On average, each video had 2,060,379 “likes” and 47,922 comments. None of the videos were duplicated and 6 did not show tics. In addition, 2 videos were removed from the TikTok webpage before all reviewers could assign a Likert score. Therefore, we included 94 videos for the assessment of tic phenomenology and 92 videos for Likert scale scoring.

### Phenomenology

Table 1 shows the frequencies of the portrayed tic-like phenomenology and table 2 presents illustrative examples. Many videos showed a very high number of different tics in a single day (>10 tics; 52.1%). Coprophenomena was frequently portrayed (copropraxia: 20.2%; coprolalia: 53.2%). Context-dependent coprophenomena and coprolalia consisting of long phrases (>3 words) were the most common atypical characteristics noted. Many subjects reacted positively to coprophenomena and used it for a comedic effect. In some videos, coprophenomena was directed at a specific person. Some subjects replaced words in a sentence or song lyric with obscene words while keeping the same number of syllables and/or rhythm of the sentence or song.

**Table 1.** Phenomenology of tic-like behavior in TikTok videos (N=94).

|  | **n (%)** |
| --- | --- |
| More than 10 different tics in a single day | 49 (52.1%) |
| **Motor tics** | 92 (97.9%) |
| Area of the body involved |  |
| Face and neck | 88 (93.6%) |
| Rest of the body | 78 (83.0%) |
| Motor tics disrupt a voluntary action | 36 (38.3%) |
| Copropraxia | 19 (20.2%) |
| Prolonged or sustained copropraxia (> 3 seconds) | 1 (5.3%)* |
| Context-dependent copropraxia | 4 (21.1%)* |
| High number of different types of copropraxia (>3) | 0 (0.0%)* |
| Subject reacts positively to copropraxia | 3 (15.8%)* |
| Echopraxia | 2 (2.1%) |
| Palipraxia | 0 (0.0%) |
| Context-dependent motor tics | 51 (54.3%) |
| Aggression to others | 18 (19.1%) |
| Throwing objects | 21 (22.3%) |
| Self-injurious behaviors | 26 (27.7%) |
| Prolonged or sustained motor tics (> 3 seconds) | 4 (4.3%) |
| Tics causing falls | 6 (6.4%) |
| **Phonic Tics** | 83 (88.3%) |
| Phonic tics disrupt speech | 8 (8.5%) |
| Coprolalia | 50 (53.2%) |
| Coprolalia consisting of long phrases (>3 words) | 20 (40.0%)* |
| Context-dependent coprolalia | 25 (50.0%)* |
| High number of different types of coprolalia (>3) | 6 (12.0%)* |
| Subject reacts positively to coprolalia | 11 (22.0%)* |
| Echolalia | 5 (5.3%) |
| Palilalia | 3 (3.2%) |
| Context-dependent phonic tics | 51 (54.3%) |
| Long phrases (>3 words) | 43 (45.7%) |

\* Percentages reported are based on the total videos of the subcategory.

**Table 2.** Illustrative examples of tic-like behavior in TikTok.

| Examples | Additional atypical features* |
| --- | --- |
| **Coprophenomena** |  |
| Saying “Fuck off, you bitch”. | Long phrases (>3 words) |
| Saying “dumb fuck” instead of “dim sum” while cooking dim sums. | Replaces words for coprolalia of similar syllables. |
| Replacing parts of the lyrics of the “Bob the Builder” cartoon theme song for “Fuck off right now!” | Context-dependent, long phrases (>3 words), follows the tune of the song. |
| Saying “You chunky ass bitch” directed to a woman. | Context-dependent, long phrases (>3 words), directed to a specific person. |
| **Context-dependent** |  |
| Saying “Simba” while smearing a substance on own forehead, then saying “Mufasa” while smearing the same substance on another person’s forehead. The subject appears frustrated. | Non-stereotyped, complex, refers to a popular character (in the Lion King movie, a fruit is smeared on Simba’s forehead. Mufasa is Simba’s dad), ego-dystonic. |
| Saying “Mix it with your glasses” while stirring a mixture of slime using the subject’s glasses. Then saying “wear them again” while putting the glasses back without cleaning them. The subject appears frustrated. | Non-stereotyped, complex, subject narrates the ongoing motor behavior, ego-dystonic. |
| Interrupting a person that says: “Believe it or not (...) I’m...” by saying “I’m gay. I knew it!”. | Non-stereotyped, long phrase (>3 words), follows context of the conversation. |
| Responding to a person that asks: “Do you know what is really annoying?” by saying “You” and then apologizing. | Follows the context of the conversation, ego-dystonic. |
| Saying: “Ew stop. My ears are bleeding” as a response to someone else’s singing and then apologizing. | Follows context of the conversation, long phrase (>3 words), ego-dystonic |
| Elbowing, head butting, and karate-chopping a Jenga tower in a non-stereotyped matter while producing several vocalizations. The subject loses | Non-stereotyped, complex, ego-dystonic. |
| the game as a result. |  |
| Saying: “Ejaculate on the board” while pouring a white sauce in a cutting board. The subject appears frustrated. | Non-stereotyped, complex, long phrase (>3 words), ego-dystonic. |
| Saying: “It’s a hat” while putting a hot pan lid on own head causing pain. | Non-stereotyped, complex, self-injurious behavior, ego-dystonic. |
| Changing multiple parts of the lyrics of the nursery rhyme Old McDonald. For example, changing “Everywhere a moo-moo” for “Everywhere a BigMac” or adding “called Peppa” after “and on his farm he had a pig”. Subject appears frustrated. | Non-stereotyped, follows the tune of the song, refers to popular characters (Peppa is a cartoon pig), ego-dystonic. |
| **Aggression to others** |  |
| Slapping someone’s arm while saying “don’t touch me” followed by an apology, then screaming “I said don’t!” followed by a second apology. The other person was doing the subject’s hair. | Non-stereotyped, second phrase refers to first phrase. |
| Slapping another person repeatedly in a non-stereotyped matter for a few seconds and then returning to regular activities. | Non-stereotyped. |
| **Self-Injurious Behavior** |  |
| Putting makeup, slime, or other non-edible or injurious objects inside the mouth or on the tongue. | Non-stereotyped, context-dependent. |
| Hitting and slapping different parts of the body in a non-stereotype matter for a few seconds and then continuing with regular activities. | Non-stereotyped |
| Jaw locked causing to bite lips for ~30 minutes resulting in injuries to the lips. | Prolonged motor tic (>3 seconds). |
| **Throwing objects** |  |
| Throwing different objects like eggs, milk, or flour while cooking or baking. | Non-stereotyped, context-dependent. |
| Performing a “challenge” consisting of holding an egg without dropping it for as long as possible. Often the challenge ends with the subject throwing the egg after only a few seconds. | Context-dependent. |
| **Long Phrases** |  |
| Saying “That’s not a tic” or “That’s not a tic, I just dislike you” after an offensive tic-like behavior. | Context-dependent, phrase refers to a previous tic-like behavior. |
| Saying: “I bite men” followed by “Thank god you are not a man” while pointing at someone. | Context-dependent, second phrase refers to first phrase. |

\* Some features are not included in Table 1 because they were difficult to assess systematically in all videos. However, we included them as qualitative observations in some videos.

CDT were also frequently portrayed (motor: 54.3%; phonic: 54.3%). Commonly, CDT were complex actions and resulted in frustration or embarrassment (e.g., were ego-dystonic). Phonic CDT were often portrayed as unwanted, occasionally insulting, verbal responses that were consistent with the context of an ongoing conversation. Other common patterns of CDT include complex motor actions while verbally describing such action, or referring to popular characters (e.g, mentioning a popular farm animal cartoon while cooking that specific type of meat). In extreme cases, videos showed subjects almost constantly reacting involuntarily to the environment, giving the appearance of a complete loss of inhibition.

Aggression toward other people or objects, present in 19.1% of videos, was often non-stereotyped and, occasionally, accompanied by complex verbal behavior. Similarly, self-injurious behavior (SIB), present in 27.7% of videos, was commonly non-stereotyped. Putting non-edible or injurious material inside the mouth or on the tongue was seen frequently. Throwing objects, present in 22.3% of videos, was commonly seen in the context of cooking or baking. A “challenge” consisting of holding an egg for as long as possible was portrayed by multiple subjects. Often, the challenge ended with the subject throwing the egg after a few seconds. Long phrases (>3 words) were commonly seen (45.7% of videos) and were frequently context-dependent. In some cases, tic-like behavior consisting of prolonged phrases referenced a previous behavior, such as denying that the previous behavior was a tic. We found one example of a motor tonic tic that was portrayed as lasting ∼30 minutes causing injuries to the subject.

We qualitatively noted that tic-like behaviors more frequently involved the trunk and extremities, but most videos also showed tic-like behaviors involving the head and neck. Since our analysis is based on entire videos and not individual tic-like behaviors, the predominance of behaviors involving the trunk and extremities is not apparent in table 1. We did not extract the complexity of tic-like behaviors due to difficulty in assessing it accurately in all videos. However, we noted that many videos showed a mixture of simple and complex tic-like behavior with a preponderance of complex behaviors.

### Likert score

The primary reviewers’ median Likert scale score, after averaging both primary reviewers, was 4.5 (IQR:4-5) and the senior reviewers’ median Likert score was 5 (IQR: 3-5). The Spearman correlation coefficient between primary and senior reviewers was 0.46 (p<0.001; Figure 1).

**Figure 1.**
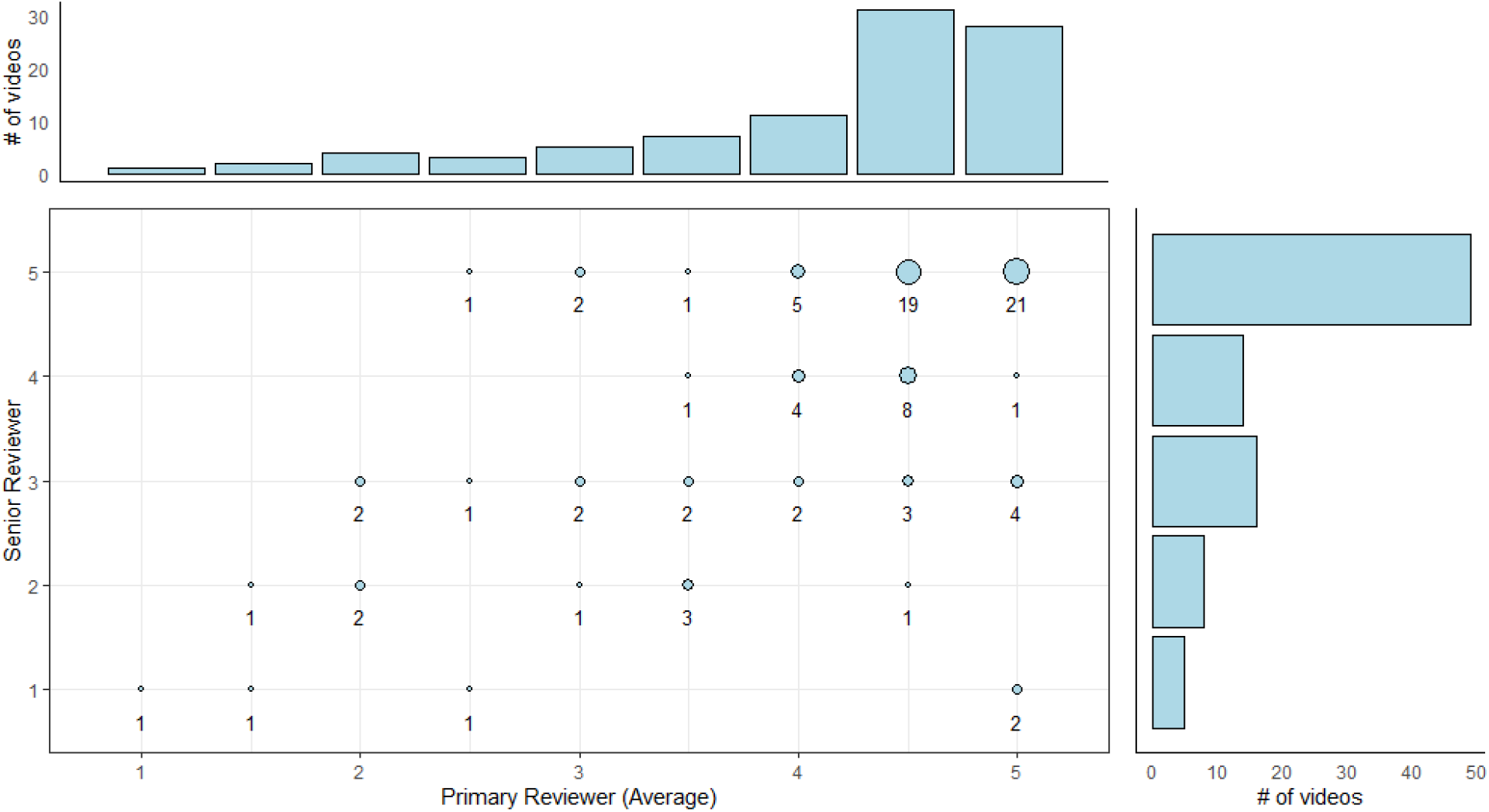
Scatter plot of the Likert scale scores between senior reviewers (y-axis) and primary reviewers (x-axis). The Likert scores from primary reviewers was averaged. The size of the circles represents the number of videos at each point. The bar plots for senior and primary reviewers are shown in the corresponding margins.

## DISCUSSION

In this assessment of TikTok videos indexed in March 2021 by “#tourettes”, we found a high frequency of movements, vocalizations, and severe behavioral disturbances more characteristic of functional tic-like behaviors than of tics. Both primary and senior reviewers judged most videos as poorly consistent with a primary tic disorder (Likert score 4-5). Importantly, this video review is not equivalent to a clinical evaluation of the individuals in these videos. A clinical in-person assessment would include many elements not obtainable solely through video review. However, it does show that these videos could be misleading to the general public.

Coprophenomena, seen in ∼15% of TS patients,^19^ were overrepresented in these videos (copropraxia: 20.2%; coprolalia: 53.2%) and often had atypical features. Coprophenomena portrayed in these videos was frequently context-dependent, and included long phrases. Although urges to make insulting remarks about a person’s trait can be seen in TS^20^, coprophenomena in TS generally consists of short words or movements presenting out of context and without clear triggers.^3^ CDT were also very frequent (motor: 54.3%; phonic: 54.3%). Tic-like behavior that is very consistent with the situation of the subject is more frequently seen with FTLD than TS.^21^ Tics triggered by an external stimulus are present in ∼20% of TS patients but, unlike tics portrayed in these videos, they are more commonly simple and ego-syntonic.^16^

Rage attacks, which might include physical aggression, are present in 20-67% of patients with TS^22^, and SIB are seen in 35% of TS patients.^23^ However, these behaviors are not always tics and can also be caused by obsessive-compulsive thoughts, mood disorders, or impulsivity.^22-26^ Additionally, aggression to others due to different psychiatric disorders, including functional and factitious disorder, can be misdiagnosed as TS.^27^ We found a high number of videos showing aggression to others (19.1%) and SIB (27.7%) with atypical features for a tic. Although we were unable to define the exact cause of these behaviors, most videos implied or explicitly said that these were tics. This is particularly worrisome giving the modeling phenomenon that has been reported in recent FTLD patients.^8^

Other atypical phenomenology noted in the videos includes a very high number of different tics, preponderance of complex behaviors or behaviors involving the trunk and extremities, tic-like behaviors consisting of long phrases, and tic-like behaviors consisting of throwing objects. In TS, simple tics are more common than complex tics,^28-30^ and tics affecting the eyes, face, and neck are more common than tics affecting the rest of the body.^31^ Although phonic tics can include phrases,^30^ these are generally short. In our experience, tics consisting of throwing objects are very rare but we couldn’t find systematic evaluations of this feature.

A recent single-center analysis of TS TikTok videos was conducted to assess phenomenology. ^32^ Interestingly, the authors extracted videos the same week that we did therefore they likely analyzed similar sets of videos as the current study. They also found high rates of coprophenomena, complex tics, and self-injurious behaviors. In addition to these findings, our study added further details about atypical phenomenology and included overall impression of videos a using Likert scale by reviewers from multiple centers. Considering both studies, there is mounting evidence that multiple specialists from different centers, including adult and child neurologists, have come to similar conclusions about the accuracy of TS TikTok videos.

The present study was prompted by the authors’ clinical experience with an unexplained surge in adolescents presenting to our clinics with functional tic-like behaviors. The atypical phenomenology in the videos we reviewed has been frequently reported in patients with FTLD during the COVID-19 pandemic.^7-9^ Some patients present with a tic that is frequently seen in TikTok but otherwise would be rarely seen^7, 9^ and some unusual tics were shared by different subjects in these videos.^32^ All authors have noticed the same trends in our clinical practice.

In several recent studies, others have suggested a possible causal relation between TikTok TS videos and FTLD. ^6-8, 10^ One hypothesis is that the rise in FTLD is caused by “social contagion” or modeling.^8, 12^ While to date we do not have strong evidence to support this hypothesis, disease modeling is seen in functional neurologic disorders, and modeling of tics portrayed in social media has been reported in a previous case of mass functional (psychogenic) illness.^33, 34^ Alternatively, since some of the teenagers diagnosed with FTLD had a history of mild tics in childhood, echophenomena in genetically susceptible patients has also been postulated to explain some cases.^7^ Although infectious or post-infectious etiologies can present fulminantly, a recent review of movement disorders associated with proven COVID-19 infection found no cases of tics or tic-like events.^35^

In the treatment of Tourette Syndrome and tic-like behaviors, it is recommended to minimize attention (negative or positive) on tics or tic-like behaviors.^36, 37^ Increased attention to the abnormal behaviors can result in exacerbation and perpetuation of symptoms in both disorders but is more prominent in FTLD.^2^ These videos often show excessive attention to the person with tics. This effect can also extend to viewers reinforcing abnormal behaviors. We recommend that FTLD patients avoid watching TS TikTok videos. Additionally, the effect of these videos in TS patients has not been studied.

Our study has several limitations. First, behaviors due to obsessive-compulsive symptoms, disinhibition, or impulsivity can resemble tics and may have been mischaracterized by the reviewers. However, the included events were portrayed as tics and are likely to be interpreted as such by a lay audience. Second, the categorization of tics based on short videos is imperfect.^38, 39^ To limit subjectivity, multiple reviewers evaluated each video.

Finally, among several factors we could not assess by video and which therefore raise questions for future studies, we wish to suggest the possible role of secondary gain as a contributor to the high prevalence of severe, atypical features. A previous study found that negative portrayals of TS are more popular in social media.^18^ We note the possibility that the high number of views, comments, and likes for these videos may have resulted in psychological and/or economic benefits to the individuals posting the videos. While many individuals in these videos express an interest in increasing TS awareness, the present analysis suggests a risk of creating a highly inaccurate perception of TS. This problem may exist for other disorders, as a recent study reported that some TikTok videos intended to create eating disorders awareness could be interpreted as portraying pro-eating disorders messages.^17^

## CONCLUSION

In sum, current TikTok videos are poorly representative of TS and could be misleading to the general public. The detailed description in this paper is intended to help pediatricians, child psychiatrists, and child neurologists recognize this atypical phenomenology to guide additional inquiry about social media exposure and treatment. Videos of tics are a notable example of the proliferation of misleading videos portraying medical conditions during the pandemic. However, similar trends have been reported in other neurologic and psychiatric disorders.^17, 40^ Although our findings could suggest an association between TikTok videos and the current spike in FTLD, our study was not designed to evaluate this. Further research is needed to define the relationship between social media and FTLD.

## Data Availability

Data is available at request.

## Abbreviations

TS: Tourette Syndrome
FTLD: Functional tic-like disorder
CDT: Context-dependent tics
SIB: Self-injurious behavior

