## Supplementary Materials for "The phenomenology of tics and tic-like behavior in TikTok"

**PREDEFINED TIC PHENOMENOLOGY AND SELECTED DEFINITIONS**

- Does the video show movements or sounds that are portrayed as tics or could be interpreted to represent tics by a lay audience?
- Does the video show more than 10 different tics (both vocal and phonic) in a single day?

**Motor tics**

- Areas of the body.
  - Select if the motor tics involve the “Face and Neck” or “Rest of Body”. If both areas are involved in the video, report both as present.
- Motor tics disrupt an ongoing voluntary action.
  - Tics must clearly disrupt an ongoing voluntary action. Tics that briefly interrupt voluntary movements are not sufficient to consider this item as being present.
- Copropraxia.
  - Prolonged or sustained copropraxia.
    - This item refers to a single sustained obscene gesture or complex behavior for more than 3 seconds. Repetitive obscene gestures going on for longer than 3 seconds are not sufficient to consider this item as being present.
- Context-dependent copropraxia.
  - This item refers to obscene actions that are strongly influenced by the environment. This includes obscene actions triggered by an external stimuli OR obscene actions that are very specific to the subject situation of the subject.
  - Do not consider this item present if the only manifestation of context-dependent copropraxia echopraxia where the subject copies someone else’s obscene gesture.
- High number of different types of copropraxia.
- This item refers to more than 3 different obscene gestures or actions that are portrayed on the same day. This item should not be considered to be present if the video shows multiple different obscene gestures or actions that are portrayed on different days.
- Subject reacts positively to copropraxia.
- This item refers to the immediate response of the subject to an obscene action. Consider this item to be present if the subject seems amused or pleased right after the obscene action. Do not consider this item to be present if the subject is indifferent, embarrassed, or upset immediately after an obscene action.
- Echopraxia.
- Palipraxia.
- Context-dependent motor tics.
- This item refers to actions that are strongly influenced by the environment. This includes actions triggered by an external stimuli OR actions that are very specific to the subject situation of the subject.
- Do not consider this item present if the only manifestation of context-dependent motor tics is echopraxia or palipraxia.
- Aggression to others.
- Throwing objects.
- Self-injurious behaviors.
- Prolonged and sustained motor tics.
- This item refers to a sustained tonic action or complex behavior lasting more than 3 seconds. Repetitive simple behaviors. Repetitive short actions going on for longer than 3 seconds are not sufficient to consider this item as being present.
- Tics causing falls.

**Phonic tics**

- Phonic tics disrupt speech
- Tics must clearly disrupt an ongoing sentence causing the subject to lose the train of thought. Tics that briefly interrupt speech are not sufficient to consider this item as being present.
- Coprolalia
- Coprolalia consisting of long phrases.
- This item refers to obscene phrases of more than 3 words. Repeating an obscene word or short phrase (3 words or less) multiple times in a row is not sufficient to consider this item as being present.
- Context-dependent coprolalia.
- This item refers to obscene words or phrases that are strongly influenced by the environment. This includes obscene words or phrases triggered by an external stimuli OR obscene words or phrases that are very specific to the subject situation of the subject.
- Do not consider this item present if the only manifestation of context-dependent coprolalia is echolalia where the subject copies someone else’s obscene word or phrase.
- High number of different types of coprolalia.
- This item refers to more than 3 different obscene words or phrases that are portrayed on the same day. This item should not be considered to be present if the video shows multiple different obscene words or phrases that are portrayed on different days.
- Subject reacts positively to coprolalia.
- This item refers to the immediate response of the subject to an obscene word or phrase. Consider this item to be present if the subject seems amused or pleased right after the obscene word or phrase. Do not consider this item to be present if the subject is indifferent, embarrassed, or upset immediately after an obscene word or phrase.
- Echolalia
- Palilalia
- Context-dependent phonic tics
- This item refers to words or phrases that are strongly influenced by the environment. This includes words or phrases triggered by an external stimuli OR words or phrases that are very specific to the subject situation of the subject.
- Do not consider this item present if the only manifestation of context-dependent phonic tics is echolalia or palilalia.
- Long phrases
- This item refers to a tic consisting of long phrases (>3 words. Repeating a word or short phrase (3 words or less) multiple times in a row is not sufficient to consider this item as being present.

**Likert scale**

Considering the limitation of short and edited videos, rate each video form 1-5:

- 1= All of the movements or sounds portrayed are typical of primary tic disorders such as Tourette syndrome.
- 2= The majority of the movements or sounds portrayed are typical of primary tic disorders such as Tourette syndrome.
- 3= Some of the movements or sounds portrayed are typical of primary tic disorders such as Tourette syndrome, but some are not typical of these disorders.
- 4= The majority of the movements or sounds portrayed are not typical of primary tic disorders such as Tourette syndrome.
- 5= None of the movements or sounds portrayed are typical of primary tic disorders such as Tourette syndrome.
